# Cost, cost-effectiveness and budget impact analysis of near point-of-care GeneXpert testing for STIs in South Africa: leveraging current capacity to address high prevalence of *Chlamydia trachomatis*, *Neisseria gonorrhoeae* and *Trichomonas vaginalis*

**DOI:** 10.1101/2024.12.13.24319006

**Authors:** Nkgomeleng Lekodeba, Katherine Snyman, Brooke E Nichols, Lise Jamieson

**Affiliations:** Health Economics and Epidemiology Research Office, Faculty of Health Sciences, University of the Witwatersrand, Johannesburg, South Africa; Department of Global Health and Development, London School of Hygiene and Tropical Medicine, London, United Kingdom; Department of Global Health, Boston University School of Public Health, Boston, Massachusetts, USA; Department of Global Health, Amsterdam Institute for Global Health and Development, Amsterdam UMC, University of Amsterdam, Amsterdam, Netherlands; South African Department of Science and Innovation/National Research Foundation Centre of Excellence in Epidemiological Modelling and Analysis (SACEMA), Stellenbosch University, Stellenbosch, South Africa

**Keywords:** Sexually transmitted infections, Point-of-care, GeneXpert testing, cost-effectiveness, Adolescent girls and young women, Incremental cost-effectiveness ratios

## Abstract

**Background:** South Africa has high sexually transmitted infections (STIs) prevalence and currently implement syndromic management, which has limitations such as untreated asymptomatic infections and antibiotic misuse. Diagnostic tools, like GeneXpert may offer potential improvements. We evaluated costs, cost-effectiveness and budget impact of reallocating GeneXpert capacity for STIs testing.

**Methods:** We developed static analytical model using previously collected data. Over one-year time horizon from provider perspective, we compared costs reported in 2024 USD and outcomes of syndromic management and nine scenarios using near point-of-care GeneXpert testing for *Neisseria gonorrhoeae*, *Chlamydia trachomatis*, and *Trichomonas vaginalis* in various target groups, including symptomatic individuals, antenatal care (ANC) attendees and HIV testers (Adolescent girls and young women (AGWY), and adults). Univariate sensitivity analysis was conducted to assess uncertainty of key parameters.

**Results:** Cost per person treated and correctly treated ranged from $21-$29 (syndromic management) and $88-$579 in GeneXpert scenarios. Syndromic management cost the healthcare system an estimated $24 million, GeneXpert testing would cost substantially more: $207 million (symptomatic), $116 million (ANC attendees), $1.7 billion (HIV testers), and $310-$884 million for targeted/combined approaches involving ANC attendees, AGWY and symptomatic individuals, and increase number of cases correctly treated by over 3-fold. Of scenarios modelled, two were cost-effective: 1) AGYW HIV testers and adults with STI symptoms, and 2) adults (15-49 years) having either HIV test or STI symptoms, incremental cost per additional case correctly treated was $515 and $1,079, respectively. While they are cost-effective, they would cost $2.26 and $10.52 billion over 5-years, respectively, compared to $145 million in syndromic management. Cost of cartridge was most influential parameter in sensitivity analysis.

**Conclusions:** Prioritizing symptomatic individuals, high-risk groups (i.e HIV testers), and cost-effective interventions can improve cases correctly treated but requires additional budget. These findings support the need for targeted strategies to optimise clinical and economic benefits of GeneXpert testing for STIs.

## Introduction

Worldwide, four curable sexually transmitted infections (STIs) – syphilis (*Treponema pallidum*), *Neisseria gonorrhoeae* (NG), *Chlamydia trachomatis* (CT), and *Trichomonas vaginalis* (TV) – account for over 1 million new infections daily, including, annually, 156.3 million new cases of trichomoniasis, 128.5 million new cases of chlamydia, 82.4 million new cases of gonorrhoea, with the majority occurring in low- and middle-income countries (LMIC) (1). Sub-Saharan Africa (SSA) has a disproportionately high prevalence of these infections the highest age-standardized incidence rates and a greatest number of disability-adjusted life years lost (2), driven by limited access to healthcare, diagnostics, and prevention, as well as socio-economic factors like poverty and gender inequality, which hinder treatment uptake and increase exposure(3,4). South Africa has one of the largest burdens of curable STIs in the world with the prevalence notably higher among adolescent girls and young women (AGYW) and in pregnant women, where reported prevalence of STIS can reach up to 40% (5–11). In addition, South Africa has the largest population of people living with HIV (PLHIV) in the world, with about 8 million PLHIV in 2024 (12). Co-infection with HIV is common, and people living with HIV are more vulnerable to STI infection, further compounding the public health challenge (10). Untreated STIs can lead to severe health consequences, including infertility, pelvic inflammatory disease, ectopic pregnancies, stillbirths, and an increased risk of HIV acquisition and transmission, making them a critical public health concern (13). Gonorrhoea’s growing antimicrobial resistance (AMR) further complicates STI control, highlighting the urgency of addressing this issue (14–16). While STIs are preventable, and most are treatable or curable, effective case management and prevention strategies, such as point-of-care diagnostics and clinical treatment protocols, are essential for breaking the transmission cycle.

In South Africa and throughout SSA, syndromic management is the standard approach for curable STIs (17), but it has limitations, including the high prevalence of untreated asymptomatic infections (18) and risks of overtreatment and antibiotic misuse (19). The WHO recommends etiological testing for curable STIs to enhance diagnostic accuracy and improve disease management, especially in LMICs (20). While syphilis rapid tests are widely used for antenatal screening and are considered highly cost-effective (21–23), no rapid tests are currently available or used in South Africa for other curable STIs, such as NG, CT, and TV. In LMICs, where screening for these STIs does occur, Nucleic Acid Amplification Tests (NAATs) or Polymerase Chain Reaction (PCR) testing are the most common methods, but they are often prohibitively expensive (24,25). GeneXpert (Cepheid, Sunnyvale, CA, USA) is a PCR-based test that enables point-of-care (POC) testing for NG, CT, and TV, with options for both off-site and on-site testing. Studies in South Africa (26) and Botswana (27) reported incremental cost-effectiveness ratios (ICERs) for various GeneXpert strategies ranging from $93 to $5,445 compared to syndromic management. While evidence on the cost-effectiveness of POC versus off-site testing for curable STIs in SSA remains limited, current findings suggest that off-site testing is more cost-effective (24).

Recent studies from South Africa show that off-site GeneXpert testing for *Neisseria gonorrhoeae* (NG) and *Chlamydia trachomatis* (CT) is both less cost-effective (26) and more-costly (28) compared to on-site testing using GeneXpert. The GeneXpert diagnostic platform has been in use in South Africa since 2011 as part of an extensive molecular diagnostic program for tuberculosis (TB) (29). Currently, the program is expanding to incorporate multiple manufacturers, moving beyond reliance on a single platform. Between April 2022 and March 2023, the National Health Laboratory Service (NHLS) conducted approximately 2.57 million Xpert MTB/RIF Ultra tests nationwide, which is below the current capacity of 9.7 million tests per year (30). As part of this expansion, it is anticipated that some of the excess GeneXpert capacity will become available for testing of other infections such as CT, NG and TV (31,32).

In this study, we assessed the potential impact of reallocating excess GeneXpert capacity to test for STIs with overlapping symptomology, focusing on both cost and health outcomes including the number of cases correctly and incorrectly treated. We then evaluated the cost, cost-effectiveness and budget impact of current syndromic management and multiple near-POC GeneXpert testing scenarios for individuals seeking care for STIs in South Africa.

## Materials and Methods

### Model structure and approach

We developed a Microsoft Excel-based static decision analytical mathematical model to estimate cost and cost-effectiveness of multiple STIs screening strategies, and assessed the budget impact of near-POC GeneXpert testing for STIs diagnostics (Fig 1). The base case represented the current syndromic management of STIs, which considers a population of adults aged 15-49 years presenting with vaginal discharge syndrome (VDS) and male urethritis syndrome (MUS). We constructed multiple hypothetical scenarios which comprised population-specific scenarios, targeted and/or combination scenarios that implemented near-POC GeneXpert platforms to screen and test for STIs, including NG/CT and/or TV. For each scenario, we estimated health outcomes (total number of cases treated, cases correctly treated and excess antibiotic use) and costs (total health system costs, cost per case treated and correctly treated). We then used the information to conduct the cost-effectiveness and budget impact analysis.

**Fig 1.**
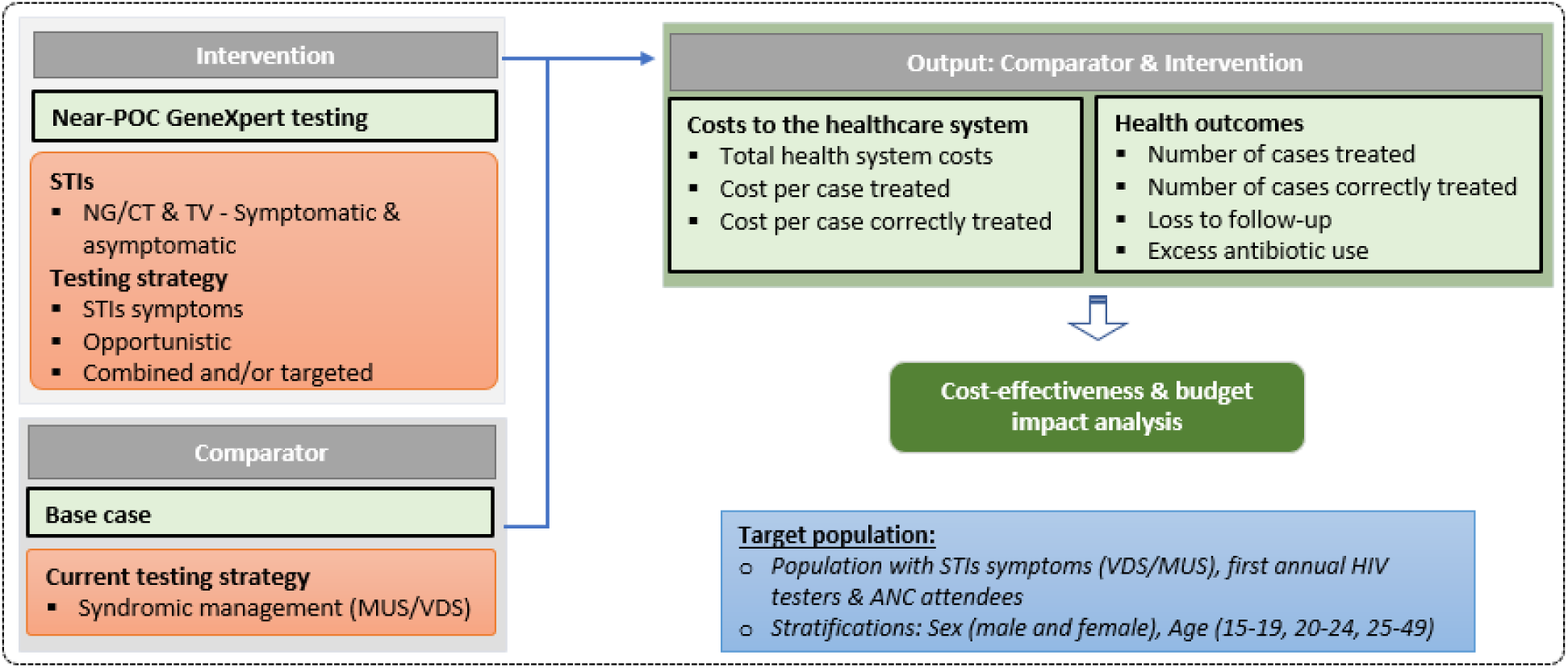
The modelling structure and approach*. *Abbreviations: STIs=sexual transmitted infections, ANC=antenatal care; MUS=male urethritis syndrome, VDS=vaginal discharge syndrome, HIV=human immunodeficiency virus; NG=neisseria gonorrhoea; CT=chlamydia trachomatis; TV=trichomonas vaginalis; POC = point-of-care

### Modelled scenarios

We modelled near-POC GeneXpert testing scenarios in three broad categories: 1) near-POC syndromic GeneXpert testing – a set of scenarios testing populations presenting with MUS and VDS; 2) near-POC opportunistic GeneXpert testing – testing those presenting for their first annual HIV test, or pregnant women attending antenatal care; 3) Combined and/or targeted near-POC GeneXpert testing scenarios – a selection of scenarios that were either targeted to specific sub-populations or combined any of the previously mentioned scenarios. A description of base case and modelled scenarios is shown in Table 1.

**Table 1:**
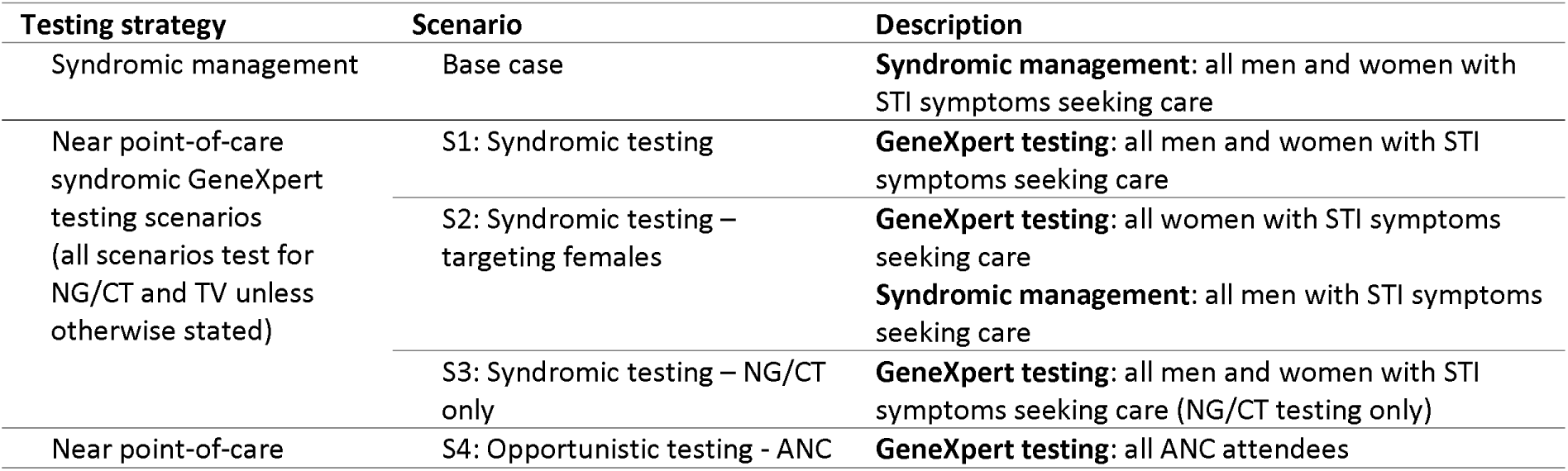
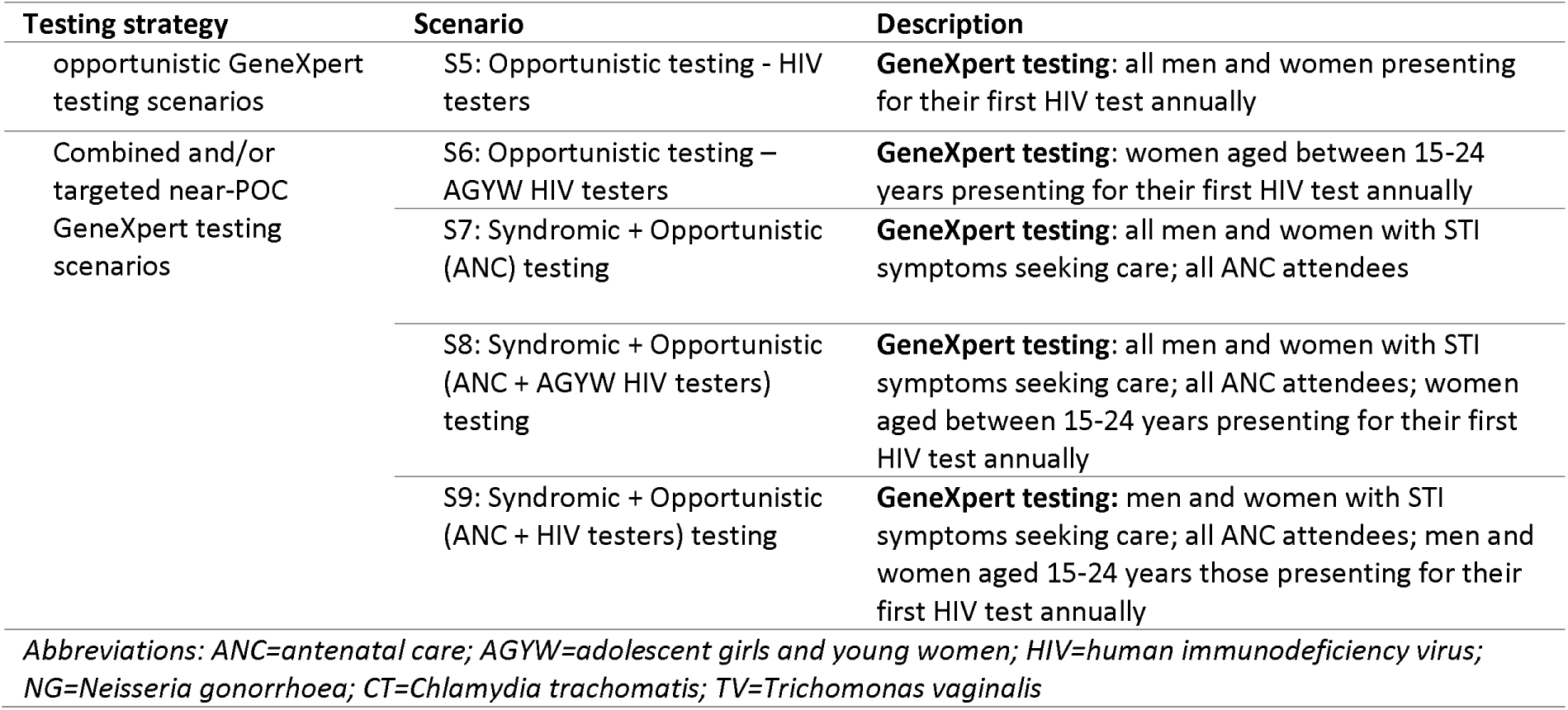
Summary of modelled scenarios.

### Input parameters

To inform the parameters for the model, we performed a scoping review of the literature which evaluated GeneXpert for STI diagnosis in South Africa and throughout SSA for *Chlamydia trachomatis, Neisseria gonorrhoea* and *trichomonas vaginalis*. We report key model input parameters in Table 2 and our literature search strategy in S1 Table. Briefly, recent studies conducted in South Africa provided age- and sex-stratified prevalence for VDS and MUS (33) and for NG, CT and TV (11)and the co-infection rates of NG and CT (28) (Table 2). For simplicity and considering studies from South Africa found low co-infection rates with TV, we did not include these in our model (8–10). Prevalence of STIs in women attending ANC were determined by pooling estimates separately for HIV positive pregnant women (5,10) and HIV negative pregnant women (7,10), which we incorporated that into our model accordingly. We obtained the sensitivity and specificity of VDS/MUS (5) for predicting actual STI prevalence, and the sex-stratified sensitivity and specificity of GeneXpert testing for NG, CT (34) and TV (35). To inform the size of the target populations across different scenarios, we used several sources. Age- and sex-stratified South African population data was obtained from Statistics South Africa (12),and the projected number of HIV testers was obtained from the Thembisa model (36). For the main analysis, we used estimates from 2024 for both target populations. For antenatal care attendees, we used the annual number of live births (37) adjusted for the percentage of singleton births (38) and ANC coverage at public health facilities (39).

**Table 2:** Key model input parameters.

| Parameter | Male | Female | Source |
| --- | --- | --- | --- |
| **Population parameters** |  |  |  |
| Population size |  |  | (12) |
| 15-19 years | 2,757,213 | 2,729,231 |  |
| 20-24 years | 2,369,402 | 2,347,230 |  |
| 25-49 years | 12,065,426 | 12,067,638 |  |
| HIV testers (number of people) |  |  | (36) |
| 15-19 years | 456,803 | 932,464 |  |
| 20-24 years | 807,048 | 1,352,708 |  |
| 25-49 years | 3,766,765 | 3,915,335 |  |
| **Antenatal care attendees** |  |  | (37) |
| 15-49 years | N/A | 595,691 |  |
| **Sensitivity and specificity** |  |  |  |
| VDS/MUS sensitivity | 91.5% | 44.9% | (5) |
| VDS/MUS specificity | 60.3% | 74.2% |  |
| GeneXpert sensitivity, CT | 93.0% | 91.0% | (34) |
| GeneXpert specificity, CT | 100.0% | 100.0% |  |
| GeneXpert sensitivity, NG | 96.0% | 93.0% |  |
| GeneXpert specificity, NG | 100.0% | 100.0% |  |
| GeneXpert sensitivity, TV | 89.6% | 97.8% | (35) |
| GeneXpert specificity, TV | 99.3% | 99.4% |  |
| **Epidemiological parameters** |  |  |  |
| **Prevalence of MUS/VDS in PHC attendees** |  |  | (33) |
| 15-19 years | 1.9% | 2.2% |  |
| 20-24 years | 7.3% | 5.7% |  |
| 25-49 years | 3.7% | 2.5% |  |
| **Prevalence of CT in PHC attendees** |  |  |  |
| 15-19 years | 4.7% | 14.7% | (11) |
| 20-24 years | 8.3% | 15.0% |  |
| 25-49 years | 4.4% | 5.6% |  |
| **Prevalence of NG in PHC attendees** |  |  |  |
| 15-19 years | 1.7% | 4.5% |  |
| 20-24 years | 2.6% | 6.1% |  |
| 25-49 years | 1.8% | 2.6% |  |
| **Prevalence of TV in PHC attendees** |  |  |  |
| 15-19 years | 0.9% | 11.2% |  |
| 20-24 years | 1.7% | 15.5% |  |
| 25-49 years | 6.6% | 17.3% |  |
| **Proportion all NG and CT cases that are NG/CT co-infections** | 14.0% | 14.0% | (28) |
| **% lost to follow-up after diagnostic test** | 8.0% | 8.0% | (40) |
| **% of live births that are singletons** | N/A | 98.5% | (38) |
| **% ANC coverage** | N/A | 94.0% | (39) |
| **% of pregnant women seeking antenatal care** | N/A | 70.0% |  |
| **HIV prevalence in ANC attendees** | N/A | 27.5% | (41) |
| **Prevalence of NG in ANC attendees** | N/A | 5.5% (HIV+); 3.1% (HIV-) | Pooled analysis |
| **Prevalence of CT in ANC attendees** | N/A | 18.3% (HIV+); 20.4% (HIV-) | HIV +: (10) |
| **Prevalence of TV in ANC attendees** | N/A | 15.1% (HIV+); 10.0% (HIV-) | HIV -: (7,10) |
| **Cost and resource utilization parameters (all costs reported in 2024 USD), applies to males and females** |  |  |  |
| Standard patient consultation cost | \$19.95 | | (42) |
| % of PHC visit cost for an asymptomatic case | 50% |  | Assumed |
| Number of additional time to draw sample for GeneXpert testing | 10 minutes |  | Assumed |
| Transportation costs to laboratory per sample | \$0.11 | | NHLS personal communication |
| Registration costs at laboratory per sample | \$1.02 | | (43) |
| MTB/XDR GeneXpert testing at centralized laboratory | \$54.84 | | |
| MTB/XDR GeneXpert cartridge | \$14.90 | | (44) |
| NG/CT GeneXpert cartridge | \$16.20 | | |
| TV GeneXpert cartridge | \$19.00 | | |
| Collection swab for GeneXpert | \$1.52 | | |
| % overhead cost at laboratory | 10% |  | (28) |
| % additional cost for External Quality Assessment at laboratory | 4% |  | Assumed |

Under the base-case syndromic management scenario, we assumed that all primary healthcare (PHC) attendees presenting with STI symptoms and seeking care would receive and adhere to treatment as per the *Standard Treatment Guidelines*. These guidelines outline a syndromic approach, which “treats the signs or symptoms (syndrome) of a group of diseases rather than targeting a specific disease.” This approach enables the simultaneous treatment of one or more conditions that often co-occur and has been established as the preferred management strategy (17). Our analysis focused on VDS and MUS, as these clinical symptoms are most commonly associated with NG, CT and TV. While the guidelines also include lower abdominal pain (LAP) in women as a syndrome associated with these infections, we excluded it from our model due to insufficient data and to maintain simplicity in the model design (17). Under all near-POC GeneXpert testing scenarios, different populations were targeted as described in detail in Table 1, including those with STI symptoms, ANC attendees and HIV testers. We further assumed that since clients need to return for a next appointment to get their results and treatment, they were at risk of loss-to-follow-up at a rate of 8%, across all populations in these scenarios (40).

#### Cost analysis

We conducted our analysis from the provider perspective, representing the South African Government. Costs were initially converted to South African Rand (ZAR) in the year of purchase, subsequently inflated to 2024 ZAR using the consumer price index and converted to United States Dollar (USD) using the January-June 2024 mean exchange rate (1 USD = 18.7 ZAR), with all costs presented in 2024 USD (45,46).

##### Clinic-related costs

For syndromic management, we estimated the cost of a clinic consultation using a recent PHC costing analysis, and used the average visit cost of staff, equipment, consumables, and overheads (42). We assumed one visit per client, which included triage, screening/diagnosis and treatment. For near-POC GeneXpert testing scenarios, we assumed that clients with STI symptoms incurred the full cost of a clinic consultation visit, while asymptomatic clients incurred half the cost. For sample collection, we included the cost of a GeneXpert collection swab and an additional 10 minutes of consultation time with a professional nurse at a mid-point pay grade (47). As results would need to be delivered at an additional visit, we assumed this additional cost at the same rate as a standard patient consultation. Furthermore, we assumed one day of training for three professional nurses per facility per year, ensuring that at least one trained nurse is always available in facility and that all nurses will be trained over five years (48).

##### Laboratory-related costs

Diagnostic costs encompassed expenses related to transportation, sample registration, equipment, human resources, supplies and materials, overheads, and external quality assessment (EQA) testing. For testing scenarios which included the use of cartridges for both TV and NG/CT testing, laboratory costs (excluding cartridges) were assumed to double. Staff, equipment, and consumable costs were based on a recent costing analysis of MTB/XDR GeneXpert testing conducted in a high-throughput centralized laboratory within the Gauteng Province (43). We obtained the costs of the GeneXpert cartridges and swabs from the manufacturer (44). We annualized equipment costs assuming a 5-year working life and a discount rate of 4% for the annual equivalent cost (43). To adjust for test-specific consumables, the cost of the MTB/XDR GeneXpert cartridge was subtracted and replaced with the corresponding NG/CT and TV cartridge costs, as applicable. We have provided the detailed cost input parameters in Table 2.

##### Treatment costs

Treatment regimens were primarily based on the most recent *South African STI guidelines* (17) but updated to incorporate new recommendations from the 2024 WHO guidelines (49). We estimated drug costs using the weighted average contract price (weights were quantity awarded in the government tender) as recorded in the Current Master Health Product List (50), details of treatment regimens and their corresponding costs are provided in the S3 Table.

#### Cost-effectiveness analysis

To determine the final list of scenarios that will remain on the cost-effectiveness frontier, we first ranked scenarios on total health system costs of from lowest to highest. We identified and eliminated scenarios that were dominated. A scenario was considered dominated if it resulted in higher total costs to the healthcare system and fewer number of cases correctly treated to the next highest ranked scenario. A ICER per additional case correctly treated for each scenario was then calculated by dividing the difference in total health system costs (incremental cost) by the difference in cases correctly treated (incremental effect) of one scenario compared to the next best scenario on the cost-effective frontier. We compared the ICER for each scenario with the ICER of the next best scenario, and those that were weakly dominated were identified and eliminated. A scenario was considered weakly dominated if it resulted in smaller effect (lower number of cases correctly treated), but had a higher ICER compared to the next highest ranked scenario.

#### Sensitivity analysis

We conducted one-way deterministic sensitivity analysis by varying model input parameters to identify parameters that were most influential to the ICER per person correctly treated in affecting the cost per case correctly treated under the base case scenario and ICER per case correctly treated for the scenarios on the cost-effectiveness frontier. We conducted a univariate deterministic sensitivity analysis by varying key input costs parameters diagnostic costs (cost of cartridges and EQA), training cost and other input parameters (sensitivity of syndromic management and near-POC GeneXpert scenarios).

#### Budget impact analysis

We estimated budget impact for the current syndromic management (base case) and scenarios on the cost-effectiveness frontier (S5 and S6) over a 5-year period. The 5-year period was chosen to align with the South African government’s medium-term planning which occurs over a period of 5 years. For base case and each scenario on the cost-effectiveness frontier, we adjusted for input parameters – South African population (12), HIV testers (36) and ANC attendees (37) annual projections and cost parameters.

### Ethics statement

This study did not require ethics approval because it involved publicly available and previously published data.

## Results

### Epidemiological health outcomes

The analysis included a total population of 34.3 million male and females aged 15-49 years for the current syndromic management and different GeneXpert testing scenarios. In the base case and near-POC GeneXpert testing (S1-S3), 1.16 million symptomatic individuals were physically examined and tested through near-POC GeneXpert testing platforms, with varying population undergoing physical examinations and near-POC GeneXpert testing, targeted by sex and STIs (Table 3). Near point-of-care opportunistic GeneXpert testing scenarios (S4 and S5) included 595,691 ANC attendees and 11.2 million HIV testers among the overall population. Targeted and/or combined near point-of-care GeneXpert testing scenarios testing (S6) included 2.3 million AGYW and 1.07 million male and females of other age groups presenting with symptoms went through syndromic management. Other targeted and/or combined near point-of-care GeneXpert testing scenarios (S7-S9) expanded coverage, testing 1.73 million (S7), 3.92 million (S8), and 4.88 million (S9) through GeneXpert. A detailed total population included in the base case and all scenario is shown in S2 Figure.

**Table 3:**
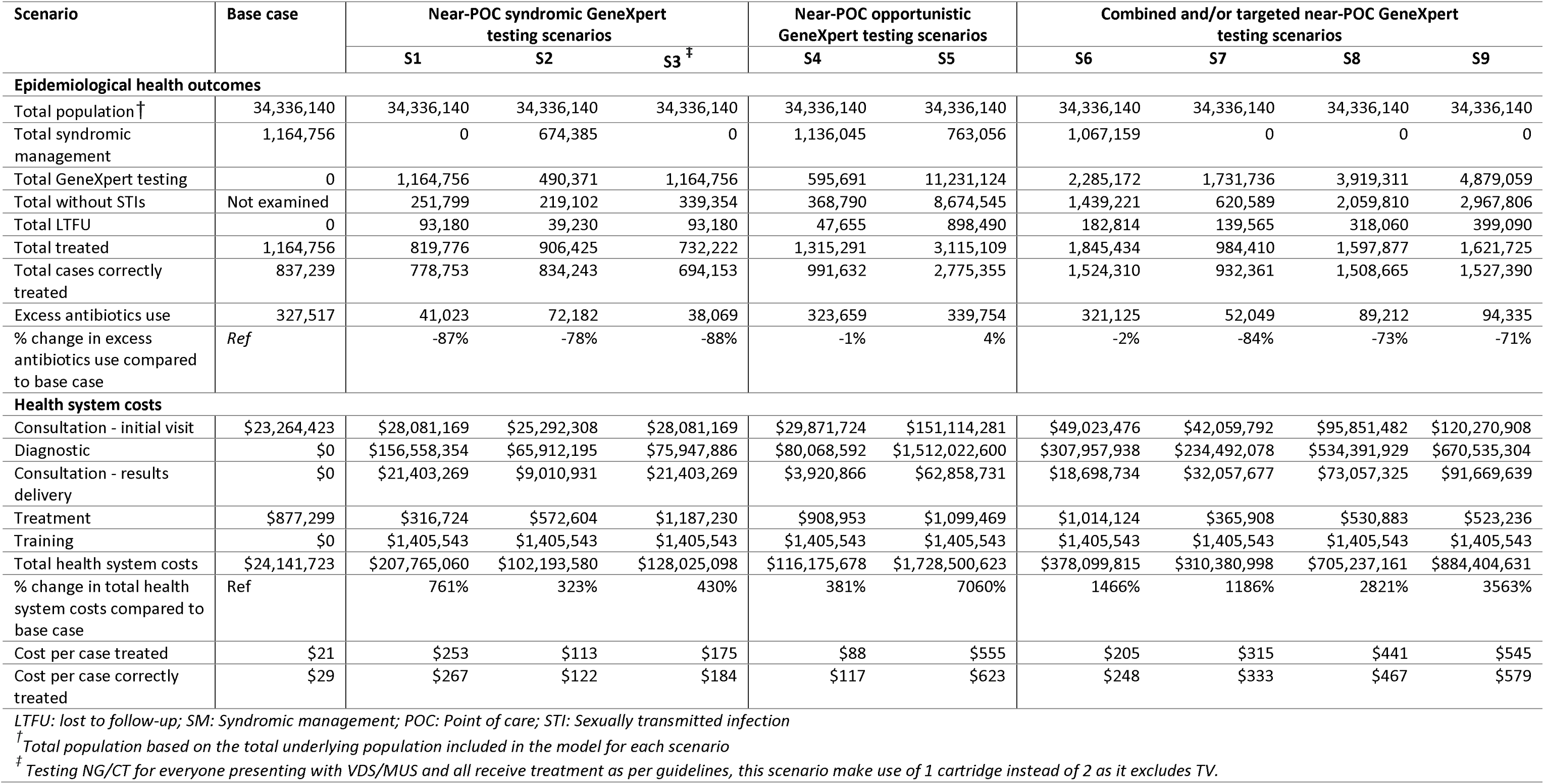
Epidemiological health outcomes and total cost, cost per person treated and cost per person correctly treated (2024 USD), by scenario.

In the base case, all individuals presenting with symptoms were treated for STIs, with 837,239 cases correctly treated, leading to 327,517 cases of excess antibiotic use. In the near-POC syndromic GeneXpert testing scenarios (S1–S3), the number of correctly treated cases was lower due to targeted testing (women only), limited STI testing (NG and CT), and LTFU up to 93,180 cases. In the near-POC opportunistic GeneXpert testing scenarios, 1.3 million (S4) and 3.1 million (S5) individuals were treated and of these 992,000 (S4) and 2.78 million (S5) cases correctly treated. In the targeted and combined near-POC GeneXpert testing scenarios, testing AGYW HIV testers (S6) resulted in 1.8 million individuals treated, with 1.5 million correctly treated. Testing adults with STI symptoms and ANC attendees (S7) included 984,410 individuals treated and 932,361 cases correctly treated. Scenarios S8 and S9, with the highest testing coverage, had up to 1.6 million individuals treated and 1.5 million cases correctly treated, and LTFU ranging between 318,060 to 399,090 (Table 3). Compared to the base case, S5 was the only scenario that resulted in an increase in excess ABs use, followed by lower reduction among S4 and S6 of up to 4% but higher in other GeneXpert testing scenarios (up to 88%).

### Cost outcomes

In the base case, the cost per patient visit, excluding treatment, was $20, primarily driven by staff-related costs (S4 Table). Implementing near-POC GeneXpert testing for NG/CT and TV increased costs to $166 and $178 per symptomatic and asymptomatic individual (excluding treatment costs) and limiting testing to NG/CT reduced costs to $97.60 and $110, respectively. Diagnostic costs were the main cost drivers, accounting for up to75% to 80% for symptomatic and asymptomatic NG/CT and TV testing, and between 60% to 67% of cost for when testing only asymptomatic and symptomatic NG/CT.

Total costs, cost per person treated and cost per person correctly treated were lowest in the base case scenario (Table 3). Across GeneXpert testing scenarios, the cost per person treated ranged from $88-$545, and cost per person correctly treated $117-$623. The main cost drivers for the GeneXpert testing scenarios were diagnostic costs (up to 87% of total cost) and staff costs (up to 39% of total cost) all other costs account for less than 1% of total costs. While the base case cost the healthcare system $24 million, implementing GeneXpert syndromic testing would cost the healthcare system up to $207 million (+761% more than base case), near-POC opportunistic GeneXpert testing scenarios increase in cost up to $1.7 billion (+7060%) and targeted and/or combined near-POC GeneXpert testing scenarios cost the healthcare system up to $884 million (+3563%).

### Cumulative cost and number of GeneXpert tests

To assess the costs and feasibility of scaling up testing strategies, we visualized cumulative costs and GeneXpert tests performed across all GeneXpert testing scenarios (Fig 2). The size of bubble represents the cumulative number of cases correctly treated and numbers above or attached to each bubble represent number of cases correctly treated. Each scenario progressively expands the testing population, with each patient requiring two tests: one for NG/CT and one for TV. The dotted vertical line indicates the current annual national testing capacity of 9.7 million tests. We found that all scenarios, except scenarios 5 (S5) – (Opportunistic testing - HIV testers), and scenario 9 (S9) – (Syndromic + Opportunistic testing: ANC attendees and HIV testers), are feasible to implement within the current capacity.

**Fig 2.**
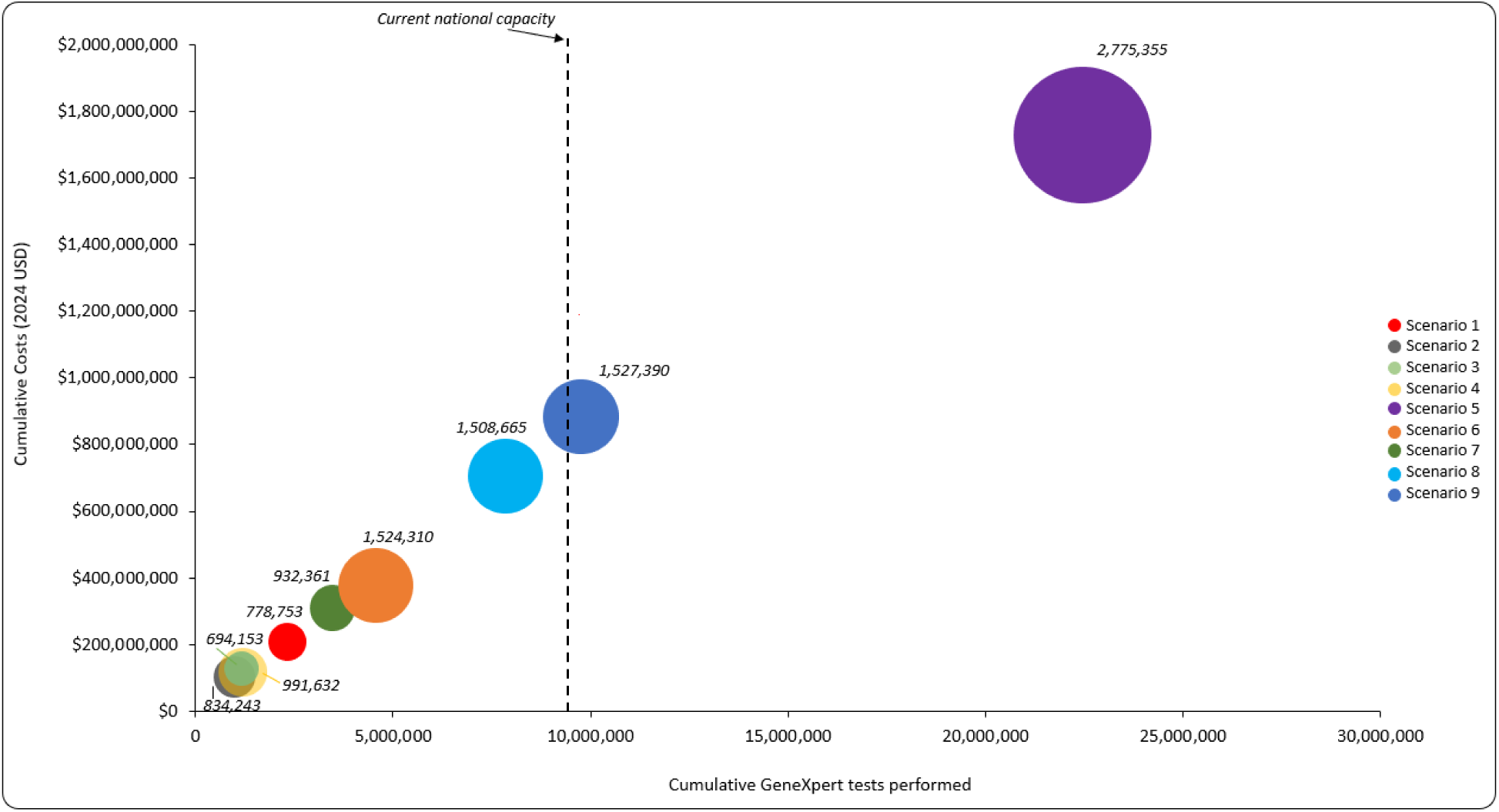
Cumulative cost and number of GeneXpert tests performed by scenario*. \**See table 1 for the description of each scenario*

### Cost-effectiveness

The results of all scenario analysed are included in the Table 4, and of those, only two scenarios were on the cost-effectiveness frontier. All of these scenarios on the cost-effectiveness frontier resulted in an increase in total health system costs and the number of cases correctly treated. This include opportunistic near-POC GeneXpert testing (S5) and targeted and/or combined near-POC GeneXpert testing scenario (S6). The ICER per additional case correctly treated ranged between $515 to $1,079 (Table 4, Fig 3). Scenarios that are not on the cost-effectiveness frontier were regarded as dominated or weakly dominated due to resulting in higher total health system costs and fewer number of cases correctly treated compared to the next best scenario.

**Fig 3.**
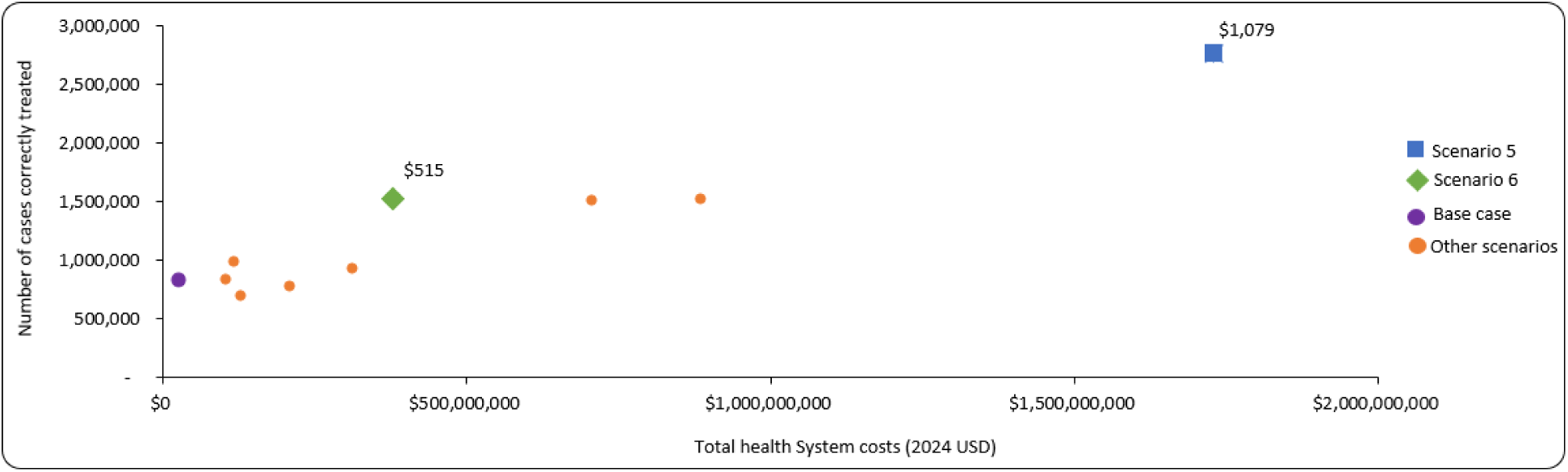
Total cost, total number of cases correctly treated, ICER, and cost-effectiveness of scenarios that increase number of cases correctly treated.

**Table 4:** Total costs, total number of cases correctly treated, ICER and cost-effectiveness of scenarios that increase number of cases correctly treated.

| Scenario and descriptions<sup>†</sup> | Total costs, 2024<br>USD (% change<br>compared to base<br>case) | Total number of<br>cases correctly<br>treated (%<br>change compared<br>to base case) | ICER per<br>additional case<br>correctly treated<br>(USD) <sup>‡</sup> |
| --- | --- | --- | --- |
| **Base case:** Syndromic management: adults aged 15-49 years with STI symptoms seeking STIs care | \$24,141,723 (n/a) | 837,239<br>(n/a) | - |
| **S2:** GeneXpert testing: all women with STI symptoms seeking care, and syndromic management: all men with STI symptoms seeking care | \$102,193,580 | 834,243 | Dominated |
| **S4:** GeneXpert testing: all ANC attendees | \$116,175,678 | 991,632 | Weakly dominated |
| **S3:** GeneXpert testing: all men and women with STI symptoms seeking care (NG/CT testing only) | \$128,025,098 | 694,153 | Dominated |
| **S1:** GeneXpert testing: all men and women with STI symptoms seeking care | \$207,765,060 | 778,753 | Dominated |
| **S7:** GeneXpert testing: all men and women with STI symptoms seeking care; all ANC attendees | \$310,380,998 | 932,361 | Dominated |
| **S6:** GeneXpert testing: women aged 15-24 years presenting for their first HIV test annually and adults aged 15-49 years with STI symptoms seeking STIs care | \$378,099,815<br>(+1466%) | 1,524,310<br>(+82%) | \$515 (compared to base case) |
| **S8:** GeneXpert testing: all men and women with STI symptoms seeking care; all ANC attendees; women aged between 15-24 years presenting for their first HIV test annually | \$705,237,161 | 1,508,665 | Dominated |
| **S9:** GeneXpert testing: men and women with STI symptoms seeking care; all ANC attendees; men and women aged 15-24 years those presenting for their first HIV test annually | \$884,404,631 | 1,527,390 | Weakly dominated |

| Scenario and descriptions<sup>†</sup> | Total costs, 2024 USD (% change compared to base case) | Total number of cases correctly treated (% change compared to base case) | ICER per additional case correctly treated (USD)<sup>‡</sup> |
| --- | --- | --- | --- |
| S5: GeneXpert testing: adults aged 15-49 presenting for their first HIV test annually and adults aged 15-49 years with STI symptoms seeking STIs care | \$1,728,674,520 (+7060%) | 2,775,355 (+231%) | \$1,079 |

<sup>†</sup>Scenarios were ordered in ascending order of increasing total health system costs
<sup>‡</sup>ICERs per additional case correctly treated were calculated by comparing scenarios to the previous scenarios that was on the cost-effective frontier; see details in methods section.

Near-POC GeneXpert testing in women aged 15-24 years presenting for their first HIV test annually and all syndromic male and females aged 15-49 with STIs symptoms seeking STIs care (S6), resulted in the increase in number of cases correctly treated by 82% and a 16-fold increase in total health system costs. compared to base case, S6 resulted in ICER per additional case correctly treated of $515. The most expensive scenario on the cost-effectiveness frontier was near-POC GeneXpert testing for adults aged 15-49 presenting for their first HIV test annually and all syndromic male and females aged 15-49 with STIs symptoms seeking STIs care (S5) resulted in an ICER per additional case correctly treated of $1,079 per additional case correctly treated. The number of cases correctly treated increased by 231%. Implementing this scenario will require an additional budget $1.7 million compared to the base case (Table 4, Fig 3).

### Sensitivity

The sensitivity analysis shows that the cost of cartridge was the most influential input cost parameter for the ICER per case correctly treated for both scenarios (fig 4). Other parameters that were influential to the ICER per case correctly treated include sensitivity (VDS/MUS) and EQA. Staff cost, LTFU and training costs were least influential parameters to the ICER. In the base case sensitivity analysis, staff and overheads costs were the most influential cost input parameters (S5 figure).

**Fig 4.**
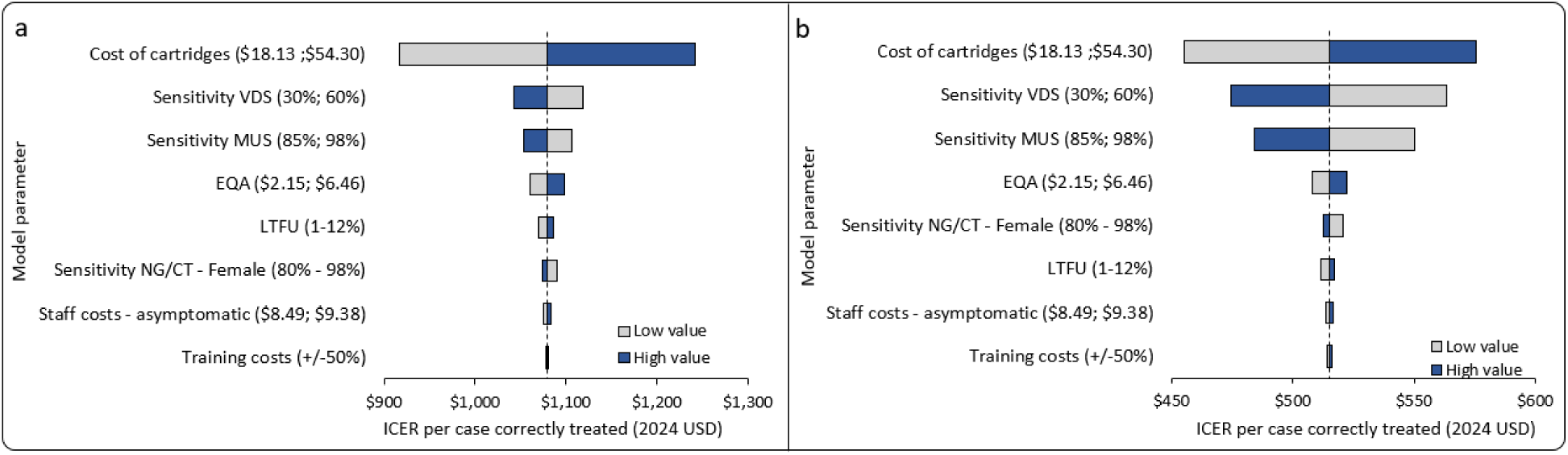
Sensitivity analysis for the ICER per case correctly treated for the scenarios on the cost-effectiveness frontier*. *The dashed line in fig 4 represent the uncertainty impact on the ICER per case correctly treated under scenario 5 (**A**) and scenario 6 (**B**).

#### Budget impact

Table 5 shows the budget impact of the current syndromic management and the scenarios on the cost-effectiveness frontier over a 5-year period (2025–2029). Under the current base case, the results show that it will cost the government 25 million to treat individuals presenting with symptoms in 2025 and which will amount to $145 million in 5-year period. In S5, it would cost the government $1.84 billion to treat HIV testers with STIs and syndromic individuals seeking STIs care and it is projected to increase by projected to cost the government $2.26 billion over the 5-year period and $399.81 in year 2025 and $507.30 in year 2029.

**Table 5.** Budget impact of the base and scenarios on the cost-effectiveness frontier.

| Year | Base case | Scenario 5 | Scenario 6 |
| --- | --- | --- | --- |
| 2025 | \$25,634,580 | \$1,843,932,546 | \$399,808,489 |
| 2026 | \$27,279,675 | \$1,964,716,262 | \$424,278,556 |
| 2027 | \$29,067,322 | \$2,095,967,094 | \$450,484,323 |
| 2028 | \$31,001,111 | \$2,235,237,204 | \$478,358,398 |
| 2029 | \$32,984,245 | \$2,382,657,792 | \$507,299,121 |
| **Total** | **\$145,966,933** | **\$10,522,510,898** | **\$2,260,228,887** |

## Discussion

In this study, we evaluated the potential impact of reallocating excess GeneXpert capacity for STI testing, focusing on both cost and health outcomes, including accurate and inaccurate treatment cases. Our findings highlight that prioritizing symptomatic individuals for testing can reduce unnecessary antibiotic use, thus providing a potential AMR risk mitigation. Based on current capacity (9.7 million tests), it would be possible to perform GeneXpert testing (NG/CT and TV) for all ANC attendees and AGYW HIV testers, with not enough capacity for all HIV testers (S5) and a combination of ANC attendees and HIV testers (S9) (30). Targeting ANC clinics emerged as a more feasible and beneficial approach for opportunistic screening compared to broader population-level interventions. Of these, near-POC GeneXpert testing would not only reduce unnecessary antibiotic use, but also correctly treat more individuals. While implementing GeneXpert testing for all individuals undergoing HIV testing may be prohibitively expensive and operationally challenging, focusing on specific high-risk groups, such as adolescents and young women, offers a more feasible and less costly alternative. These insights underscore the importance of tailored strategies to maximize both the clinical and economic impact of GeneXpert deployment for STI management.

Our estimated costs for GeneXpert diagnostics exceeded the current NHLS laboratory cost list for TB Xpert testing (R465 or $24.90 per test) (51) which also informed prior literature estimates for centralized NG/CT testing ($28.58 per case diagnosed) (28). This discrepancy is likely explained by the NHLS subsidizing MTB/XDR Xpert assays. A similar subsidy for STI GeneXpert testing could reduce costs, suggesting that our estimates—and resulting ICERs—may be overestimated. Smith et al. reported comparable costs per patient diagnosed with on-site GeneXpert testing ($80.90), though this included microscopy and represented true POC, i.e. POC testing that is based at the facility and test results are provided within the patient’s time at the facility, rather than near-POC testing (26). Nonetheless, incremental costs for GeneXpert diagnostics remain high. Several studies have identified strategies to reduce costs for NG/CT etiological testing, such as using in-house PCR testing (52), pooling samples (53), conducting GeneXpert testing in centralized laboratories (28), and targeting screening based on risk factors (54). A mixed approach in Botswana for pregnant women, involving POC at high-volume sites and centralized laboratories elsewhere, resulted in lower costs per case averted compared to each of these strategies on their own or syndromic management (27).

Our analysis was limited to the primary outcome of correctly treated cases, without considering potential downstream effects of correctly, or incorrectly, managing STIs. For example, the analysis could have been expanded to include broader health outcomes, such as disability-adjusted life years (DALYs), the number of congenital illnesses prevented, or other STI-related complications avoided. Including DALYs would have allowed us to compare our incremental cost-effectiveness ratios against relevant cost-effectiveness thresholds. However, our focus was on the narrower question of South Africa’s current GeneXpert capacity, associated costs, and reductions in excess antibiotic treatments. In this regard, a limitation is the aggregation of all excess antibiotic treatments, despite stronger evidence of resistance for some antibiotics compared to others (15,16). Our assumption of 9.7 million available tests consider the potential impact of scaling up other GeneXpert testing strategies, such as for TB or HPV. Further, we were limited by our choice of having a static model, which does not capture the transmission dynamics of STIs, including the risk of reinfection and the impact of treating sexual partners. These limitations suggest our findings may underestimate the broader public health benefits or challenges associated with STI testing strategies. Our findings underscore the role of STI screening strategies in mitigating antibiotic overuse and addressing AMR. Given the substantial economic burden of AMR, often exceeding their purchase cost (estimated cost associated with the consumption of one standard unit of antibiotics range: $0.01-0.07) (55), incorporating the economic burden of AMR through a “AMR penalty cost” could significantly reduce the ICERs of advanced screening technologies and costlier treatments with lower resistance potential (19). However, this analysis did not account for the broader economic and public health impacts of AMR because AMR-related costs specific to STIs and the South African context remain unquantified and were beyond the scope of this study. Finally, our scenarios assume that all people receiving opportunistic GeneXpert testing do not subsequently seek care at PHCs as part of the syndromic population. This assumption may lead to an underestimation of both cost and case estimates. However, as both costs and outcomes are likely underestimated in the same direction, the overall impact on ICERs may be minimal.

Our analysis focused on the public healthcare sector in South Africa and was conducted at the national level. As such, while our findings are generalizable within South Africa, their transferability to other Sub-Saharan African countries is limited. South Africa’s early adoption of GeneXpert in 2011, supported by national and donor funding, established a robust laboratory infrastructure, reflected in our cost estimates from centralized labs (29). In contrast, other SSA settings often face higher implementation costs due to infrastructure challenges, such as space and power requirements, and greater training needs (56). South Africa’s experienced NHLS workforce minimizes training expenses, unlike contexts donor funding allows its health system greater autonomy in setting priorities, unlike many SSA countries where donor influence shapes health agendas (57,58). These differences highlight the need for country-specific strategies for GeneXpert placement, considering infrastructure, workforce readiness, and financing capacity to ensure cost-effective implementation.

Our study highlights several policy implications and directions for future research. Our analysis found that overall, switching to GeneXpert testing will be very costly compared to the current strategy of syndromic management. These budgetary impacts will require a shift of resources from other health priorities to STI management. While we focused on repurposing existing GeneXpert capacity from MTB/RIF testing to NG/CT or TV testing, GeneXpert has broader diagnostic potential. Additional applications include screening for human papillomavirus (HPV), herpes simplex virus (HSV-1 and HSV-2), HIV and Hepatitis B and C viral load monitoring, and detection of methicillin-resistant Staphylococcus aureus (MRSA), Group B Streptococcus in pregnancy (59). This is particularly relevant as alternative rapid and POC diagnostics for NG are advancing, including lateral flow assays and rapid tests (60,61).

Moreover, other POC tools for STIs are under development, such as the GIFT screening tool for bacterial vaginosis (BV) (26,62) OSAM-TV for TV (63), and the BD Affirm VPIII assay for BV, TV, and *Candida* (64). Future research should investigate how alternative diagnostics compare to or complement GeneXpert to ensure cost-effective and sustainable approaches for managing sexually transmitted infections across diverse settings. Considering that cervical cancer is the leading cancer among women in South Africa (65), prioritizing research on the application of GeneXpert for HPV screening is a logical next step.

Additionally, extending cost-effectiveness models for GeneXpert testing—whether for STIs or other diseases—to incorporate longer-term health and economic outcomes would provide more comprehensive insights for policymaking and resource allocation.

## Conclusions

With ongoing challenges of current STIs syndromic management in South Africa, our study highlighted the potential impact of reallocating excess GeneXpert capacity for STI testing to enhance diagnostic accuracy and improve health outcomes in South Africa. Prioritizing symptomatic individuals and high-risk groups, such as HIV testers, can reduce unnecessary antibiotic use, addressing antimicrobial resistance challenges. These findings support the need for targeted and context-specific strategies to optimize the clinical and economic benefits of GeneXpert deployment for STI management. However, while our analysis shows improvements in health outcomes, adoption of near-point GeneXpert of care services may require considerable budgetary investment.

## Supporting information

S1 Table. Search Strategy for model parameter inputs

S2 Figure. Underlying population for the base case and each of the scenario

S3 Table. STI treatment regimens and associated costs

S4 Table. Testing scenario costs per patient (2024 USD)

S5 Figure. Sensitivity analysis of cost per case correctly treated under base case scenario

## Declaration of interests

We declare no competing interests.

## Authors contributions

NL, KS, LJ and BEN conceptualised the study. NL, KS and LJ did the literature reviews and compiled data for model parameterisation. NL and KS collected cost data. LJ oversaw data collection and parameterisation. NL developed the model and model structure. NL did the data analysis. NL, KS, and LJ had full access to all data in the study. LJ and BEN verified underlying data and assumptions. All authors contributed to reviewing and editing the manuscript for submission.

## Funding

This study was funded by FIND (FIND contract number: XC22-0197).

## Role of the funding source

The funder of the study had no role in study design, data collection, data analysis, data interpretation, or writing of the manuscript.

## Data availability statement

We acknowledge the importance of transparent and responsible data sharing to foster scientific collaboration and advance research. The data used for this study were collected from publicly available sources, previously published literature and do not contain any identifying information. The corresponding author - NL may be contacted by interested parties for further information.

## Notes

### Competing Interest Statement

The authors have declared no competing interest.

### Funding Statement

This study was funded by the Foundation for Innovative New Diagnostics (FIND).

### Summary of Updates

We updated population parameters to account for overlap, results: table 3 and 4, and figure 2 and 3, and added budget impact analysis (Table 5).

## References

1. World Health Organization. Global progress report on HIV, viral hepatitis and sexually transmitted infections, 2021. Accountability for the global health sector strategies 2016–2021: actions for impact. Geneva; 2021.

2. Zheng Y, Yu Q, Lin Y, Zhou Y, Lan L, Yang S, et al. Global burden and trends of sexually transmitted infections from 1990 to 2019: an observational trend study. Lancet Infect Dis. 2022 Apr;22(4):541–51.

3. Rowley J, Vander Hoorn S, Korenromp E, Low N, Unemo M, Abu-Raddad LJ, et al. Chlamydia, gonorrhoea, trichomoniasis and syphilis: global prevalence and incidence estimates, 2016. Bull World Health Organ. 2019 Aug 1;97(8):548–562P.

4. Wi T, Lahra MM, Ndowa F, Bala M, Dillon JAR, Ramon-Pardo P, et al. Antimicrobial resistance in Neisseria gonorrhoeae: Global surveillance and a call for international collaborative action. PLoS Med. 2017 Jul 7;14(7):e1002344.

5. Peters R, Klausner J, de Vos L, Feucht U, Medina-Marino A. Aetiological testing compared with syndromic management for sexually transmitted infections in HIV-infected pregnant women in South Africa: a non-randomised prospective cohort study. BJOG. 2021 Jul 22;128(8):1335–42.

6. Jarolimova J, Chidumwa G, Chimbindi N, Okesola N, Dreyer J, Smit T, et al. Prevalence of Curable Sexually Transmitted Infections in a Population-Representative Sample of Young Adults in a High HIV Incidence Area in South Africa. Sex Transm Dis. 2023 Dec;50(12):796–803.

7. De Voux A, Mvududu R, Happel A, Jaspan HB, Nyemba DC, Mashele N, et al. Point-of-Care Sexually Transmitted Infection Testing Improves HIV Preexposure Prophylaxis Initiation in Pregnant Women in Antenatal Care in Cape Town, South Africa, 2019 to 2021. Sex Transm Dis. 2023 Feb 1;50(2):92–7.

8. Green H, Taleghani S, Nyemba D, Myer L, Davey DJ. Partner notification and treatment for sexually transmitted infections among pregnant women in Cape Town, South Africa. Int J STD AIDS. 2020 Nov 1;31(13):1282–90.

9. Delany-Moretlwe S, Mgodi N, Bekker LG, Baeten JM, Li C, Donnell D, et al. High prevalence and incidence of gonorrhoea and chlamydia in young women eligible for HIV pre-exposure prophylaxis in South Africa and Zimbabwe: Results from the HPTN 082 trial. Sex Transm Infect. 2023 Nov 1;99(7):433–9.

10. Davey JNDC, Gomba Y, Bekker LG, Taleghani S, DiTullio DJ, Shabsovich D, et al. Prevalence and correlates of sexually transmitted infections in pregnancy in HIV-infected and- uninfected women in Cape Town, South Africa. PLoS One. 2019 Jul 1;14(7):e0218349.

11. Kharsany ABM, McKinnon LR, Lewis L, Cawood C, Khanyile D, Maseko DV, et al. Population prevalence of sexually transmitted infections in a high HIV burden district in KwaZulu-Natal, South Africa: Implications for HIV epidemic control. International Journal of Infectious Diseases [Internet]. 2020 Sep 1 [cited 2024 Oct 28];98:130–7. Available from: http://www.ijidonline.com/article/S1201971220304811/fulltext

12. Statistics South Africa. Mid-year population report, 2024 (P0302). 2024.

13. World Health Organization. Sexually transmitted infections fact sheet [Internet]. 2024 [cited 2024 Dec 2]. Available from: https://www.who.int/news-room/fact-sheets/detail/sexually-transmitted-infections-(stis)

14. Raccagni AR, Ranzenigo M, Bruzzesi E, Maci C, Castagna A, Nozza S. Neisseria gonorrhoeae Antimicrobial Resistance: The Future of Antibiotic Therapy. J Clin Med. 2023 Dec 18;12(24):7767.

15. Yakobi SH, Magibile YB, Pooe OJ. A systematic review of Neisseria gonorrhoeae drug resistance development in South Africa. Brazilian Journal of Microbiology. 2024 Jun 25;55(2):1053–63.

16. Mabaso N, Abbai NS. A review on Trichomonas vaginalis infections in women from Africa. S Afr J Infect Dis. 2021 Jun 10;36(1).

17. South Africa National Department of Health. Comprehensive STI Clinical Management Guidelines. 2021.

18. Mlisana K, Naicker N, Werner L, Roberts L, Van Loggerenberg F, Baxter C, et al. Symptomatic vaginal discharge is a poor predictor of sexually transmitted infections and genital tract inflammation in high-risk women in South Africa. Journal of Infectious Diseases. 2012 Jul 1;206(1):6–14.

19. Ayinde O, Ross JDC, Jackson L. Economic evaluation of antimicrobial resistance in curable sexually transmitted infections; a systematic review and a case study. PLoS One. 2023 Oct 1;18(10 October).

20. World Health Organization. Global health sector strategies on, respectively, HIV, viral hepatitis and sexually transmitted infections for the period 2022-2030 [Internet]. Geneva; 2022 [cited 2024 Dec 2]. Available from: https://iris.who.int/bitstream/handle/10665/360348/9789240053779-eng.pdf?sequence=1

21. Saweri OPM, Batura N, Al Adawiyah R, Causer LM, Pomat WS, Vallely AJ, et al. Economic in pregnancy in low- And middle-income countries: A systematic review. Vol. 16, PLoS ONE. Public Library of Science; 2021.

22. Zhang M, Zhang H, Hui X, Qu H, Xia J, Xu F, et al. The cost-effectiveness of syphilis screening in pregnant women: a systematic literature review. Vol. 12, Frontiers in Public Health. Frontiers Media SA; 2024.

23. Silke F, Earl L, Hsu J, Janko MM, Joffe J, Memetova A, et al. Cost-effectiveness of interventions for HIV/AIDS, malaria, syphilis, and tuberculosis in 128 countries: a meta-regression analysis. Lancet Glob Health. 2024 Jul 1;12(7):e1159–73.

24. Khumalo F, Passmore JAS, Manhanzva M, Meyer B, Duyver M, Lurie M, et al. Shifting the power: scale-up of access to point-of-care and self-testing for sexually transmitted infections in low-income and middle-income settings. Curr Opin Infect Dis. 2023 Feb;36(1):49–56.

25. Wi TE, Ndowa FJ, Ferreyra C, Kelly-Cirino C, Taylor MM, Toskin I, et al. Diagnosing sexually transmitted infections in resource-constrained settings: challenges and ways forward. J Int AIDS Soc. 2019 Aug 30;22(S6).

26. Smith E, Masson L, Passmore JAS, Sinanovic E. Cost-effectiveness analysis of different screening and diagnostic strategies for sexually transmitted infections and bacterial vaginosis in women attending primary health care facilities in Cape Town. Front Public Health. 2023 Mar 2;11.

27. Wynn A, Moucheraud C, Martin NK, Morroni C, Ramogola-Masire D, Klausner JD, et al. Bridging the Gap between Pilot and Scale-Up: A Model of Antenatal Testing for Curable Sexually Transmitted Infections from Botswana. Sex Transm Dis. 2022 Jan 1;49(1):59–66.

28. Marcus R, C P, Gill K, Smith P, Rouhani S, Mendelsohn A, et al. Acceptability, feasibility and cost of point of care testing for sexually transmitted infections among South African adolescents where syndromic management is standard of care. BMC Health Serv Res. 2023 Dec 1;23(1).

29. da Silva MP, Cassim N, Ndlovu S, Marokane PS, Radebe M, Shapiro A, et al. More Than a Decade of GeneXpert® Mycobacterium tuberculosis/Rifampicin (Ultra) Testing in South Africa: Laboratory Insights from Twenty-Three Million Tests. Diagnostics. 2023 Oct 19;13(20):3253.

30. National Health Laboratory Service. Annual Report 2022-2023 [Internet]. 2023 [cited 2024 Jun 1]. Available from: https://www.nhls.ac.za/wp-content/uploads/2023/11/241023-NHLS-Annual_Report.pdf

31. Cepheid. Xpert TV [Internet]. 2024 [cited 2024 Jun 1]. Available from: https://www.cepheid.com/en-GB/tests/blood-virology-womens-health-sexual-health/xpert-tv.html

32. Cepheid. Xpert CT/NG [Internet]. 2024 [cited 2024 Jun 1]. Available from: https://www.cepheid.com/en-GB/tests/blood-virology-womens-health-sexual-health/xpert-ct-ng.html

33. Frank D, Kufa T, Dorrell P, Kularatne R, Maithufi R, Chidarikire T, et al. Evaluation of the national clinical sentinel surveillance system for sexually transmitted infections in South Africa: Analysis of provincial and district-level data. South African Medical Journal [Internet]. 2023 Jul 5 [cited 2024 Oct 28];113(7):41–8. Available from: https://samajournals.co.za/index.php/samj/article/view/365

34. Xie TA, Liu YL, Meng RC, Liu XS, Fang KY, Deng ST, et al. Evaluation of the Diagnostic Efficacy of Xpert CT/NG for Chlamydia trachomatis and Neisseria gonorrhoeae. Biomed Res Int. 2020 Oct 8;2020:1–16.

35. Schwebke JR, Gaydos CA, Davis T, Marrazzo J, Furgerson D, Taylor SN, et al. Clinical Evaluation of the Cepheid Xpert TV Assay for Detection of Trichomonas vaginalis with Prospectively Collected Specimens from Men and Women. J Clin Microbiol. 2018 Feb;56(2).

36. Johnson LF, Dorrington R. Thembisa Version 4.7: A Model for Evaluating the Impact of HIV/AIDS in South Africa. [Internet]. 2024 [cited 2024 Jun 1]. Available from: https://thembisa.org/content/downloadPage/Thembisa4_7report

37. Statistics South Africa. Recorded Live Births 2023 (P0305) [Internet]. 2024 [cited 2024 Dec 5]. Available from: https://www.statssa.gov.za/publications/P0305/P03052023.pdf

38. Cutland CL, Sawry S, Fairlie L, Barnabas S, Frajzyngier V, Roux J Le, et al. Obstetric and neonatal outcomes in South Africa. Vaccine. 2024 Feb;42(6):1352–62.

39. South Africa National Department of Health, Statistics South Africa, South African Medical Research Council. Demographic and Health Survey 2016 Key Findings [Internet]. 2018. Available from: www.DHSprogram.com.

40. Travill D, Machemedze D, Zwane P, Mokhele T, Makhale L, Sokhela Z, et al. Same day testing and treatment for STIs in adolescents: results of a pilot randomised controlled trial in South Africa. In: 5th HIV Research for Prevention Conference [Internet]. Munich; 2024 [cited 2024 Dec 2]. Available from: https://programme2024.hivr4p.org/Abstract/Abstract/?abstractid=1553

41. Kufa-Chakezha T, Shangase N, Lombard C, Manda S, Puren A. The 2022 Antenatal HIV Sentinel Survey [Internet]. 2023 [cited 2024 Dec 12]. Available from: https://www.nicd.ac.za/wp-content/uploads/2024/01/Antenatal-survey-2022-report_National_Provincial_12Jul2023_Clean_01.pdf

42. Long LC, Girdwood S, Govender K, Meyer-Rath G, Miot J. Cost and outcomes of routine HIV care and treatment: public and private service delivery models covering low-income earners in South Africa. BMC Health Serv Res. 2023 Mar 11;23(1):240.

43. Cassim N, Omar SV, Masuku SD, Moultrie H, Stevens WS, Ismail F, et al. Xpert MTB/XDR implementation in South Africa: cost outcomes of centralised vs. decentralised approaches. IJTLD OPEN. 2024 May 14;1(5):215–22.

44. FIND. Cepheid pricing – FIND DxConnect Marketplace [Internet]. 2024 [cited 2024 Oct 28]. Available from: https://dxc-marketplace.finddx.org/pages/cepheid-accessible-pricing

45. STATSSA. CPI headline year-on-year rates [Internet]. 2024 [cited 2024 Oct 29]. Available from: https://www.statssa.gov.za/publications/P0141/CPIHistory.pdf?

46. South African Reserve Bank. Current Market Rates [Internet]. 2024 [cited 2024 Oct 29]. Available from: https://www.resbank.co.za/en/home/what-we-do/statistics/key-statistics/current-market-rates

47. South Africa Department of Public Service and Administration. Salary scales, with translation keys, for employees of salary levels 1 to 12 and those employees covered by Occupation Specific Dispensations (OSDs) [Internet]. 2024 [cited 2024 Dec 2]. Available from: https://www.dpsa.gov.za/policy-updates/nlrrm/remuneration_policy/annual_cost_of_living_adjustments/

48. South Africa National Department of Health. 2030 Human Resources for Health Strategy: Investing in the Health Workforce for Universal Health Coverage [Internet]. Pretoria; 2020 [cited 2024 Dec 5]. Available from: https://www.health.gov.za/wp-content/uploads/2023/06/2030-HRH-Strategy-Final.pdf

49. World Health Organization. Updated recommendations for the treatment of Neisseria gonorrhoeae, Chlamydia trachomatis and Treponema pallidum (syphilis), and new recommendations on syphilis testing and partner services [Internet]. 2024 [cited 2024 Nov 21]. Available from: https://www.who.int/publications/i/item/9789240090767

50. South Africa National Department of Health. Current Master Health Product List February 2024 [Internet]. 2024 Feb [cited 2024 Jun 6]. Available from: https://www.health.gov.za/tenders/

51. National Health Laboratory System. NHLS State Price List 2023/24 [Internet]. 2023 [cited 2024 Dec 5]. Available from: https://www.nhls.ac.za/

52. Dos Santos CG, Sabidó M, Leturiondo AL, De Oliveira Ferreira C, Da Cruz TP, Benzaken AS. Development, validation and testing costs of an in-house realtime PCR assay for the detection of Chlamydia trachomatis. J Med Microbiol. 2017 Mar 1;66(3):312–7.

53. Connolly S, Kilembe W, Inambao M, Visoiu AM, Sharkey T, Parker R, et al. A population-specific optimized genexpert pooling algorithm for chlamydia trachomatis and neisseria gonorrhoeae to reduce cost of molecular sexually transmitted infection screening in resource-limited settings. J Clin Microbiol. 2020 Sep 1;58(9).

54. McCartney DJ, Luppi CG, Silva RJC, De Araújo S, Bassichetto KC, Mayaud P, et al. Anorectal gonorrhoea and chlamydia among transgender women in Brazil: Prevalence and assessment of performance and cost of anorectal infection detection and management approaches. Sex Transm Infect. 2023;

55. Shrestha, P., Cooper, B.S., Coast, J. et al. Enumerating the economic cost of antimicrobial resistance per antibiotic consumed to inform the evaluation of interventions affecting their use. Antimicrob Resist Infect Control 7, 98 (2018). 10.1186/s13756-018-0384-3

56. Abdurrahman ST, Emenyonu N, Obasanya OJ, Lawson L, Dacombe R, Muhammad M, et al. The hidden costs of installing xpert machines in a tuberculosis high-burden country: experiences from Nigeria. Pan African Medical Journal. 2014;18.

57. James C, Gmeinder M, María A, Rivadeneria R, Vammalle C, Blecher M, et al. Health financing and budgeting practices for health in South Africa. OECD Journal on Budgeting. 2018;2017(3).

58. Agyei E, Kumah E. Navigating the complex terrain of healthcare systems in Sub-Saharan Africa: challenges and opportunities for progress. Discover Health Systems. 2024 Jun 20;3(1):39.

59. Cepheid. Molecular Diagnostic Testing Menu [Internet]. 2024 [cited 2024 Dec 9]. Available from: https://www.cepheid.com/en-US/tests.html

60. de Vos L, Daniels J, Gebengu A, Mazzola L, Gleeson B, Piton J, et al. Usability of a novel lateral flow assay for the point-of-care detection of Neisseria gonorrhoeae: A qualitative time-series assessment among healthcare workers in South Africa. PLoS One. 2023 Jun 1;18(6 June).

61. Peters RPH, Klausner JD, Mazzola L, Mdingi MM, Jung H, Gigi RMS, et al. Novel lateral flow assay for point-of-care detection of Neisseria gonorrhoeae infection in syndromic management settings: a cross-sectional performance evaluation. The Lancet. 2024 Feb 17;403:657–64.

62. Asare K, Andine T, Naicker N, Dorward J, Singh N, Spooner E, et al. Impact of Point-of-Care Testing on the Management of Sexually Transmitted Infections in South Africa: Evidence from the HVTN702 Human Immunodeficiency Virus Vaccine Trial. Clinical Infectious Diseases. 2023 Mar 1;76(5):881–9.

63. Garrett N, Mitchev N, Osman F, Naidoo J, Dorward J, Singh R, et al. Diagnostic accuracy of the Xpert CT/NG and OSOM Trichomonas Rapid assays for point-of-care STI testing among young women in South Africa: A cross-sectional study. BMJ Open. 2019 Feb 1;9(2).

64. Dessai F, Nyirenda M, Sebitloane M, Abbai N. Diagnostic evaluation of the BD Affirm VPIII assay as a point-of-care test for the diagnosis of bacterial vaginosis, trichomoniasis and candidiasis. Int J STD AIDS. 2020 Mar 12;31(4):303–11.

65. Statistics South Africa. Cancer in South Africa [Internet]. Pretoria; 2023 [cited 2024 Dec 11]. Available from: https://www.statssa.gov.za/publications/03-08-00/03-08-002023.pdf

