## Supplementaty appendix for "Cost, cost-effectiveness and budget impact analysis of near point-of-care GeneXpert testing for STIs in South Africa: leveraging current capacity to address high prevalence of *Chlamydia trachomatis*, *Neisseria gonorrhoeae* and *Trichomonas vaginalis*"

**Short title:** Optimising STIs screening through near-POC GeneXpert testing in South Africa

Nkgomeleng Lekodeba * ^a,^ Katherine Snyman* ^a,b^ , Brooke E Nichols^c,d^, Lise Jamieson^a,e^

a. Health Economics and Epidemiology Research Office, Faculty of Health Sciences, University of the Witwatersrand, Johannesburg, South Africa.

b. Department of Global Health and Development, London School of Hygiene and Tropical Medicine, London, United Kingdom

c. Department of Global Health, Boston University School of Public Health, Boston, Massachusetts, USA.

d. Department of Global Health, Amsterdam Institute for Global Health and Development, Amsterdam UMC, University of Amsterdam, Amsterdam, Netherlands.

e. South African Department of Science and Innovation/National Research Foundation Centre of Excellence in Epidemiological Modelling and Analysis (SACEMA), Stellenbosch University, Stellenbosch, South Africa.

### **S1 Table. Search Strategy for model parameter inputs**

| No | Searches | Results |
| --- | --- | --- |
| 1 | (Chlamydia) OR (Chlamydia trachomatis) OR (C. trachomatis) OR (chlamydiae OR (chlamydias) | 33,771 |
| 2 | (Point of Care) OR (Point-of-Care) OR (POC) OR (POCT) OR (Rapid) OR (decentrali* test*) OR (community-based test*) OR (outreach test*) OR (same-day diagnosis) OR (GeneXpert) OR (Xpert) OR (centralised test*) OR (Bedside Test) OR (NAT) OR (NATs OR NAAT OR NAATs OR Nucleic Acid Amplif* OR DNA Amplif* OR RNA Amplif* OR nucleic acid sequence based amplification OR NASBA OR nucleic acid hybridization OR nucleic acid hybridization OR nucleic acid test* OR nucleic acid based test* OR transcription‐mediated amplification OR self‐sustained sequence replication OR polymerase chain reaction OR PCR OR RT‐PCR OR RTPCR OR bDNA OR b‐DNA OR branched DNA OR branched‐chain DNA) | 2,561,872 |
| 3 | #1 AND #2 | 7,514 |
| 4 | (South Africa) | 186,988 |
| 5 | #3 AND #4 | 136 |
| 6 | Filters: from 2019 - 2024 | 59 |

1.1. Chlamydia trachomatis

1.2. Neisseria gonorrhoeae

| No | Searches | Results |
| --- | --- | --- |
| 1 | (Gonorrh*) OR (Neisseria gonorrhoeae) OR (N. gonorrhoeae) OR (Gonorrhea) OR (Gonorrhoeae) OR (Gonococcus) OR (Gonococcal) OR (Gonococcal infection) OR (Pelvic inflammatory disease) OR (Gonococcal epididymitis) | 44,132 |
| 2 | (Point of Care) OR (Point-of-Care) OR (POC) OR (POCT) OR (Rapid) OR (decentrali* test*) OR (community-based test*) OR (outreach test*) OR (same-day diagnosis) OR (GeneXpert) OR (Xpert) OR (centralised test*) OR (Bedside Test) OR (NAT) OR (NATs OR NAAT OR NAATs OR Nucleic Acid Amplif* OR DNA Amplif* OR RNA Amplif* OR nucleic acid sequence based amplification OR NASBA OR nucleic acid hybridization OR nucleic acid hybridization OR nucleic acid test* OR nucleic acid based test* OR transcription‐mediated amplification OR self‐sustained sequence replication OR polymerase chain reaction OR PCR OR RT‐PCR OR RTPCR OR bDNA OR b‐DNA OR branched DNA OR branched‐chain DNA) | 2,561,872 |
| 3 | #1 AND #2 | 4,835 |
| 4 | (South Africa) | 186,988 |
| 5 | #3 AND #4 | 170 |
| 6 | Filters: from 2019 - 2024 | 72 |

1.3. Trichomonas vaginalis (TV)

| No | Searches | Results |
| --- | --- | --- |
| 1 | (Trichomonas vaginalis) OR (Trichomoniasis) OR (Trichomonas vaginitis) OR (trichomonas OR trichomoniasis) OR (T. vaginalis) | 12,734 |
| 2 | (Point of Care) OR (Point-of-Care) OR (POC) OR (POCT) OR (Rapid) OR (decentrali* test*) OR (community-based test*) OR (outreach test*) OR (same-day diagnosis) OR (GeneXpert) OR (Xpert) OR (centralised test*) OR (Bedside Test) OR (NAT) OR (NATs OR NAAT OR NAATs OR Nucleic Acid Amplif* OR DNA Amplif* OR RNA Amplif* OR nucleic acid sequence based amplification OR NASBA OR nucleic acid hybridization OR nucleic acid hybridization OR nucleic acid test* OR nucleic acid based test* OR transcription‐mediated amplification OR self‐sustained sequence replication OR polymerase chain reaction OR PCR OR RT‐PCR OR RTPCR OR bDNA OR b‐DNA OR branched DNA OR branched‐chain DNA) | 2,561,872 |
| 3 | #1 AND #2 | 1,839 |
| 4 | (South Africa) | 186,988 |
| 5 | #3 AND #4 | 100 |
| 6 | Filters: from 2019 - 2024 | 42 |

### **S2 Figure. Underlying population for the base case and each of the scenario*^†^***

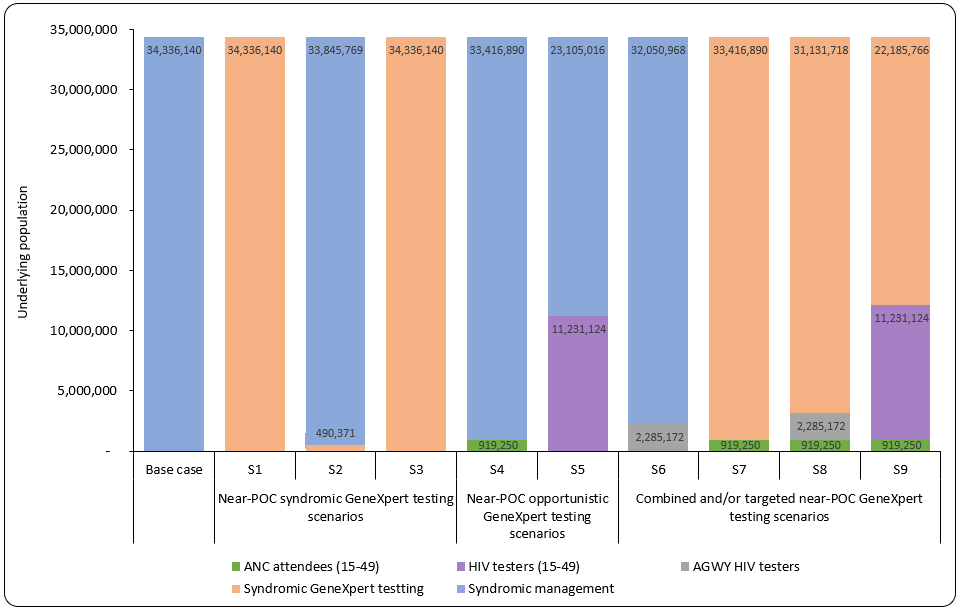

***^†^***S3 is syndromic GeneXpert testing for NG/CT only

### **S3 Table. STI treatment regimens and associated costs**

|  | Drug | Treatment course description | Cost per treatment course (2024 USD) |
| --- | --- | --- | --- |
| *Women with VDS* | |  |  |
|  | Ceftriaxone | 1g; injection; 1 Injection | 6.87 |
|  | Doxycycline | 100 mg orally, twice daily for 7 days | 7.03 |
|  | Metronidazole | 2g oral, single dose | 0.37 |
|  | Total |  | 14.27 |
| *Men with MUS, assume partner does not have VDS* | | |  |
|  | Ceftriaxone | 1g; injection; 1 Injection | 6.87 |
|  | Doxycycline | 100 mg orally, twice daily for 7 days | 7.03 |
|  | Total |  | 13.90 |
| ***N. Gonorrhoeae treatment*** | | |  |
|  | Ceftriaxone | 1g; injection; 1 Injection | 6.87 |
|  | **Total** |  | 6.87 |
| ***C. Trachomatis treatment*** | | |  |
|  | Doxycycline | 100 mg orally, twice daily for 7 days | 7.03 |
|  | **Total** |  | 7.03 |
| ***T. vaginalis treatment*** | |  |  |
|  | Metronidazole | 2g oral, single dose | 0.37 |
|  | **Total** |  | 0.37 |

### **S4 Table. Testing scenario costs per patient (2024 USD)**

|  | Syndromic management | Near-POC GeneXpert - NG/CT and TV | Near-POC GeneXpert - NG/CT only |
| --- | --- | --- | --- |
| Training Costs (total)* | N/A | 1,405,543 | 1,405,543 |
| Consultation costs per patient- symptomatic | | | |
| Assets | 1.46 | 1.46 | 1.46 |
| Overheads | 3.17 | 3.17 | 3.17 |
| Staff | 15.24 | 17.86 | 17.86 |
| Supplies | 0.09 | 1.61 | 1.61 |
| Total cost per patient | **19.96** | **24.10** | **24.10** |
| Consultation costs per patient - asymptomatic | | | |
| Assets | N/A | 0.73 | 0.73 |
| Overheads |  | 1.59 | 1.59 |
| Staff |  | 8.93 | 8.93 |
| Supplies |  | 0.80 | 0.80 |
| Total cost per patient |  | **12.05** | **12.05** |
| Diagnostics cost per patient | | | |
| Transport | N/A | 0.11 | 0.11 |
| Overheads |  | 11.83 | 5.77 |
| Equipment/ Staff/Other supplies |  | 81.86 | 40.93 |
| Cartridges |  | 36.24 | 16.68 |
| EQA |  | 4.31 | 2.10 |
| Total cost per patient |  | **134.34** | **65.59** |
| Results delivery cost per patient | | | |
| Assets | N/A | 1.46 | 1.46 |
| Overheads |  | 3.17 | 3.17 |
| Staff |  | 15.24 | 15.24 |
| Supplies |  | 0.09 | 0.09 |
| Total cost per patient |  | **19.96** | **19.96** |
| Total cost per symptomatic patient | **19.96** | **166.35** | **97.60** |
| Total cost per asymptomatic patient | **N/A** | **178.40** | **109.65** |

*Divided by number of tests for each scenario

### **S5 Figure. Sensitivity analysis of cost per case correctly treated under base case scenario**

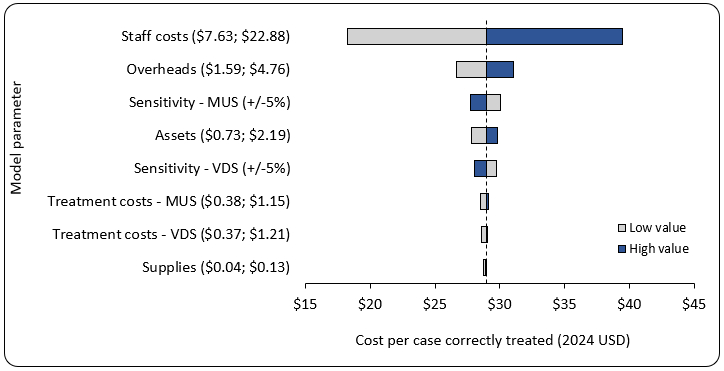
